# Quantifying the ongoing epidemic of disability after covid-19 in the UK population aged under 35 years; secondary analysis of the Covid Infection Survey

**DOI:** 10.1101/2021.08.04.21261360

**Authors:** Nicola Spiers

## Abstract

Web-based survey and anecdotal evidence suggest that many people infected with covid-19 go on to develop long covid (symptoms persisting for more than 12 weeks) despite mild initial disease. There is particular concern that this epidemic is unfolding in the younger population where vaccination rates are low. The Covid Infection Survey (CIS) has recently published estimates of the proportion of people with long covid whose daily activities are affected a little or a lot.

This paper focuses on the population aged under 35 years and uses the term ‘disabling long covid’ to describe those with symptoms lasting more than 12 weeks and daily activities limited a lot. By applying the CIS estimates to confirmed infections, with age breakdown from mid-2020 population estimates, this paper reports a first estimate of the cases of disabling long covid seeded from confirmed covid-19 infections to July 31st 2021.

Results suggest there will approximately 39,000 cases of disabling long covid in those aged under 35 seeded by Covid-19 infections confirmed to July 31st. There is a need for rapid action to prevent Covid-19 infection in the younger population and support those struggling with Long Covid-related disability.

## Introduction

Chris Whitty (CMO) and Chris Hopson (CEO, NHS Providers) have recently expressed concern about the number of young people infected with covid-19 with initially mild disease who go on to develop persistent and energy-limiting symptoms.

Much current epidemiological work on long covid (post covid-19 syndrome) is properly focused on hospitalised cohorts, who are mostly in middle or old age. But considerable anecdotal and web-based survey^1,2^ evidence points to a form of long covid that can arise from initial mild disease and last for many months, with fatigue and sleep problems as common reported symptoms. Given the ongoing rapid surge in covid-19 cases and slowdown in vaccination rates, with more than 12 million aged under 35 not double-dosed by June 30, quantifying this risk in younger people is critical.

### Measuring disabling long covid

Epidemiology of long covid to date has relied on self-reported symptoms. There is emerging consensus around the definition of long covid as persistent symptoms for 12 weeks or more. Recent data from the CIS^3^ enable a first look at the burden of disability arising from this long-term energy-limiting condition.

The CIS measures disability by a single question on limitation in daily activities, with categories not limited, limited a little and limited a lot. An advantage is that the severity and impact of the condition upon the individual and community is captured. Such questions are routinely used to quantify the population with disability in old age epidemiology. There will be differences in reporting behaviour, but the measure has meaning at population level.

For this paper, disabling long covid is defined as symptoms persistent for 12 weeks and daily activities limited a lot. These people will be struggling to stay in education and employment. They will be limited in housework and ability to support others, and maybe also limited in self-care activities. They require ongoing support from family, friends and health professionals, and without this may be at risk of developing chronic fatigue syndrome or (not the same thing but sometimes a sequel) moderate to severe depression.

## Results

The CIS^3^ estimates 962,000 individuals in the UK with long covid, including 456,000 limited a little with daily activities and 178,000 individuals of all ages with disabling long covid, in the four-week period ending 6th June 2021. This represents at least 20% of all confirmed covid-19 cases to 30th March 2021 seeding (developing into) long covid and 3.7% seeding disabling long covid, Risk of long covid increases with the risks of severe disease and hospitalisation that come with age, but should now be decreasing in the older population due to vaccination.

I focus on those aged under 35 years, as most remain unvaccinated and at risk. This is also perhaps the only way currently to begin to quantify the epidemic of initially milder, but still very unpleasant post-viral syndrome type of long covid, as opposed to the after-effects of severe initial disease, hospitalisation and ventilation. CIS estimates of numbers in June in this population are 243,000 with long covid and 28,000 (1.2% of confirmed covid-19 cases) with disabling long covid. Applying these rates to confirmed cases between April 1 and July 31, gives a further 10,900 who will go on to develop disabling long covid (Table).

**Table.** Estimated infections seeding Long Covid by age and disability

| *Infected* |  |  | *to Mar 6* |  | *Mar 7 to Jul* |  |
| --- | --- | --- | --- | --- | --- | --- |
| *Long covid* | *CIS* | *000s* | *to Jun 6* |  | *Jun 7 to Oct* |  |
|  | *estimated* |  |  |  |  |  |
|  | *%* |  |  |  |  |  |
| *Age* | *a little* | *a lot* | *a little* | *a lot* | *a little* | *a lot* |
| **<18** | 3.3 | 0.9 | 23.5 | 6.75 | 9.3 | 2.5 |
| **18-24** | 4.7 | 1.1 | 22.5 | 5.25 | 8.8 | 2.1 |
| **25 to 29** | 7 | 1.9 | 30 | 8 | 11.7 | 3.2 |
| **30 to 34** | 7.8 | 2.1 | 30 | 8 | 11.6 | 3.1 |
| **Total** | 5.3 | 1.3 | 106 | 28 | 41.4 | 10.9 |

Long Covid symptoms persisting for 12 weeks with daily activities limited a little
Disabling Long Covid persistent symptoms with daily activities limited a lot
CIS rate is estimated with Long Covid, as a fraction of confirmed Covid-19 infections

At the time of writing (August 2nd) projection of cases of COVID-19 infection for August and September is highly uncertain. If daily cases were to stabilise at close to the current rate of 20,000 per day, this would seed an estimated 8,000 further cases of disabling long covid per month. Any spike in cases such as that projected by the Imperial College roadmap out of lockdown evaluation for SAGE^4^ would add substantially more cases of disabling long covid developing by the end of 2021.

The illnesses are clearly very different, but this can be compared to the incidence of first cases of all cancers, around 30,000 per month. Even if infections stabilise, there will be many thousands of younger people struggling with disabling long covid for the foreseeable future, remembering that each will be an individual facing a very uncomfortable and challenging time.

## Limitations

REACT-2 has also released information on disabling long covid, using a directly comparable measure. I have been unable to incorporate these data as yet, but findings already published^5^ suggest that the REACT-2 estimates exceed those from the ONS infection survey. More fine-grained analysis is possible and urgently required, including work on duration of illness and vaccine protection, rapid health economic evaluation, and incorporation of disability in the SAGE projections.

As this epidemic is already upon us, my calculation is rapid and broad brush, with cautious assumptions. The CIS rates are an undercount of cases of long covid seeded by the pandemic, as many whose condition has improved or have recovered are not included. Sampling error is not large, but there will also be some non-sampling error in the CIS estimates. The numbers presented here are approximations, but are more than sufficient to indicate the magnitude of the epidemic and the possible impact of disabling long covid in the younger population.

## Conclusions

These data are new, and to date, disability is not counted in current Government pandemic projections. Against the background of the pandemic these estimates indicate the need for urgent and rapid attention to preventing disability following covid-19 in the unvaccinated population and in particular to avoid a spike in cases as schools and universities reopen.

Health service support for those suffering this long-term limiting illness will be required for the foreseeable future. Measures to prevent disabling long covid by reducing infection in the unvaccinated population should be rapidly reviewed, including encouraging vaccination uptake, restrictions on mass gatherings, mandatory face covering on public transport and in public enclosed spaces, support for those in overcrowded localities and workplaces and urgent investment in safe protocols for the return to school and university.

Disability after initially mild disease has been lost among the many challenges of the pandemic, not least because epidemiology of symptoms is complex and data on disability scarce. Allowing Covid-19 to become prevalent in the younger unvaccinated population must not be an option. It is worth remembering that a case of disabling long covid not only severely affects the individual, but also increases pressure on families and health and social care services, and contributes to labour shortage. Action on this public health crisis is overdue.

## Supporting information

coi disclosure

STROBE checklist

Excel spreadsheet with calculation detail

## Data Availability

The secondary analysis reported here used only publicly available data

## Data sharing and ethics

The secondary analysis reported here used only publicly available data and does not require ethical approval. This work is unfunded and no competing interests are declared.

## Technical Note

UK confirmed Covid-19 cases by age group were calculated by taking cumulative confirmed cases from Our World in Data to March 6th, and applying the age breakdown of confirmed cases from the Public Health England Covid-19 and flu report week 27. Estimated cases of long covid were taken from the CIS report of estimated cases for the four weeks to June 6th, linear interpolated as age groups differed.

Rates of long covid by age group were calculated by dividing numbers estimated for the four week period to June 6th by cumulative confirmed covid-19 cases to March 6th. These rates were then applied to the number of confirmed cases between March 7th and July 31st, again assuming the PHE breakdown by age and linear interpolating to give the estimated number of cases of long covid by age group seeded by infection with covid-19 during this period.

https://ourworldindata.org/explorers/coronavirus-data-explorer?zoomToSelection=true&hideControls=true&Metric=Confirmed+cases&Interval=Cumulative&Relative+to+Population=false&Align+outbreaks=false&country=~GBR

https://assets.publishing.service.gov.uk/government/uploads/system/uploads/attachment_data/file/1000373/Weekly_Flu_and_COVID-19_report_w27.pdf

https://www.ons.gov.uk/peoplepopulationandcommunity/healthandsocialcare/conditionsanddiseases/datasets/alldatarelatingtoprevalenceofongoingsymptomsfollowingcoronaviruscovid19infectionintheuk

## References

1. https://covid.joinzoe.com

2. What is long covid? 9 facts in 90 seconds. Gez Medinger 17 Nov 2020 https://www.youtube.com/watch?v=WpPnwm9UGrM/ accessed 23 June 3.

3. https://www.ons.gov.uk/peoplepopulationandcommunity/healthandsocialcare/conditionsanddiseases/datasets/alldatarelatingtoprevalenceofongoingsymptomsfollowingcoronaviruscovid19infectionintheuk

4. https://www.gov.uk/government/publications/imperial-college-london-evaluating-the-roadmap-out-of-lockdown-for-england-modelling-the-delayed-step-4-of-the-roadmap-in-the-context-of-the-delta-vpp1412July

5. O’Dowd A. Covid-19:Third of people infected have long term symptoms 24 June https://www.bmj.com/content/373/bmj.

